# Effects of genetic liability to Alzheimer’s disease on circulating metabolites across the life course

**DOI:** 10.1101/2022.03.24.22272867

**Authors:** Hannah Compton, Madeleine L Smith, Caroline Bull, Roxanna Korologou-Linden, Yoav Ben-Shlomo, Joshua A. Bell, Emma L Anderson

## Abstract

**Objective:** Alzheimer’s disease (AD) has several known genetic determinants, yet the mechanisms through which they lead to disease onset remain poorly understood. This study aims to estimate the effects of genetic liability to AD on plasma metabolites measured at seven different stages across the life course.

**Methods:** Genetic and metabolomic data from 5,648 offspring from the Avon Longitudinal Study of Parents and Children birth cohort were used. Linear regression models examined the association between higher AD liability, as measured by a genetic risk score (GRS), and plasma metabolites measured at 8, 16, 18 and 25 years of age. Two hundred twenty-nine metabolites were studied, most relating to lipid/lipoprotein traits. Two-sample Mendelian randomization was performed using summary statistics from age-stratified genome-wide association studies (GWAS) of the same metabolites for 118,466 participants from the UK Biobank, to examine the persistence of any AD liability effects into late adulthood.

**Results:** The GRS including the *APOE4* isoform demonstrated the strongest positive associations for cholesterol-related traits per doubling of genetic liability to AD, e.g., for low-density lipoprotein cholesterol (LDL-C) at age 25yrs (0.12 SD; 95% CI 0.09, 0.14), with similar magnitudes of association across age groups in ALSPAC. In the UK Biobank, the effect of AD liability decreased with age tertile for several lipid traits (e.g., LDL-C, youngest: 0.15 SD; 95% CI 0.07, 0.23, intermediate: 0.13 SD; 95% CI 0.07, 0.20, oldest: 0.10 SD; 95% CI 0.05, 0.16). Across both cohorts, the effect of AD liability on high-density lipoprotein cholesterol (HDL-C) attenuated as age increased. Fatty acid metabolites also demonstrated positive associations in both cohorts, though smaller in magnitude compared with lipid traits. Sensitivity analyses indicated that these effects were driven by the *APOE4* isoform.

**Conclusions:** These results support a profound influence of the *APOE4* isoform on circulating lipids and fatty acids from early life to later adulthood. Such lipid and fatty acid traits may be implicated in early AD pathogenesis and warrant further investigation as potential targets for preventing the onset of AD.

## INTRODUCTION

By virtue of our ageing population, the number of patients with Alzheimer’s disease (AD), the most common form of dementia, continues to rise^1^. Neuropathological hallmarks of AD precede the onset of clinical symptoms by decades^2^, yet diagnosis is often late in the disease course. Brain and cerebrospinal fluid (CSF) metabolites discriminate AD cases from controls with high accuracy^3,4^, though necessitate invasive modes of sample collection such as lumbar puncture. Therefore, great impetus remains for identification of more easily measured plasma AD biomarkers, as is beginning to be demonstrated for plasma amyloid-β^5^, which could improve our understanding of early disease aetiology.

AD involves a complex genetic architecture. Genome-wide association studies (GWAS) have illuminated many AD-associated single nucleotide polymorphisms (SNPs); the largest to date identifying 3,915 genome-wide significant variants across 38 independent loci^6^. The polymorphic *APOE* gene encodes Apolipoprotein E (ApoE). The role of the *APOE* ε4 allele (UK allele frequency 0.15^7^) in greatly elevating AD risk is unequivocal, accounting for ∼50% of total genetic susceptibility^7^, whilst the ε2 allele affords neuroprotection via mechanisms yet to be elucidated^8^. The three isoforms of *APOE* created by combinations of the ε2, ε3 and ε4 alleles, each confer differential AD risk. *APOE* functions to regulate lipid homeostasis^9^. It is hence postulated that circulating lipid perturbations are associated with both AD risk and early pathology^10^. Indeed, lipidomic studies suggest that both increased and decreased cholesterol, phospholipids, and sphingolipids^11^ may reflect neurodegeneration-associated membrane changes^12,13^. Many studies are, however, underpowered^10^ and given evaluation of AD patients in case-control studies, we cannot ascertain whether metabolic derangements are a cause or a secondary consequence of disease^13^ (i.e. biased by reverse causation), or confounded by lifestyle factors or other disease processes. The effects of medications for AD or comorbidities such as cardiovascular disease also may not be adequately accounted for^13^. Other metabolic markers such as impaired glucose homeostasis are also likely implicated in AD pathogenesis^14^, with functional brain imaging demonstrating abnormally low rates of glucose metabolism in *APOE* ε4 carriers decades before disease onset^15^. Serum amino acid profiles can discriminate AD cases from controls with high accuracy^16^, though there are inconsistencies regarding the directionality of these associations^17,18^.

In this study, we aimed to estimate the metabolic features of genetic liability to AD across the life course, with the goal of revealing early features of AD pathogenesis which may be potentially targeted to prevent the clinical onset of AD. To estimate the effect of higher liability to AD on the circulating metabolome across the life course, we constructed a genetic instrument for AD liability and examined its association with circulating metabolites measured in two studies; the Avon Longitudinal Study of Parents and Children (ALSPAC) and the UK Biobank.

## METHODS

### Study participants

ALSPAC is a population-based multi-generational birth cohort study. Eligibility of pregnant women for inclusion to ALSPAC was based on residence in a defined area of South West England and an estimated delivery date between 1^st^ April 1991 and 31^st^ December 1992^19^. Recruitment to ALSPAC occurred in four phases yielding a total of 15,454 pregnancies and 15,589 foetuses, 14,901 of whom were alive at one year^20,21^. ALSPAC offspring were eligible for this study if they had the following information recorded: genotype, sex, age, and at least one metabolic trait at any time point. A total of 5,648 individuals were eligible for analysis on at least one occasion. See Supplementary Figure 1 for full details of the eligibility criteria and Supplementary Tables 1 and 2 for descriptive statistics of the eligible ALSPAC cohort.

Ethical approval for the study was obtained from the ALSPAC Ethics and Law Committee and the Local Research Ethics Committees. Consent for biological samples has been collected in accordance with the Human Tissue Act (2004). Note that the study website contains details of all the data that is available through a searchable data dictionary and variable search tool (http://www.bristol.ac.uk/alspac/researchers/our-data/). Study data were collected and managed using REDCap electronic data capture tools hosted at the University of Bristol^24^. REDCap (Research Electronic Data Capture) is a secure, web-based software platform designed to support data capture for research studies.

Summary-level GWAS data from UK Biobank, a large-scale multicentre cohort study, were also used. Over 500,000 adults aged 40-69 years were recruited between 2006-2010 via 22 assessment centres across England Wales and Scotland. Details of the UK Biobank design, participants, quality control (QC) and its strengths and limitations have been detailed previously^22–24^.

### Assessment of genetic liability to AD

In ALSPAC, genotype was assessed using the Illumina HumanHap550 quad chip, with imputation performed with the Haplotype Reference Consortium (HRC) panel. AD susceptibility was defined using SNPs associated with AD at genome-wide significance (p≤5×10^−8^) reported by the Kunkle et al. GWAS^25^. This GWAS reports 25 loci of genome-wide significance using data from 46 case-control studies included in four AD consortia that comprise the International Genomics of Alzheimer’s Project (IGAP). A total of 21,982 cases and 41,944 cognitively normal controls were analysed, all of whom were adults of non-Hispanic white ethnicity. Amongst cases, 61.3% were female and the mean age of onset of AD was 72.9 years. Amongst controls, 57% were female and the mean age of examination was 72.4 years. Proxy SNPs that were in LD with the sentinel SNP at r^2^≥0.5 were also considered. Summary statistics for the Kunkle et al. meta-analysis are available at: https://www.niagads.org/datasets/ng00075.

Data harmonisation constituted identical coding of effect (risk-increasing) alleles in both the AD GWAS and ALSPAC datasets. If a SNP associated with AD at genome-wide significance was not present in the ALSPAC dataset, then a proxy SNP in high linkage disequilibrium (LD) (within 10,000kb, r^2^=0.8) with the AD SNP was included instead. As such, rs9331896 (associated with the CLU gene) was replaced with rs2279590. AD-associated SNPs were combined into two genetic risk scores (GRS), one including and one excluding the SNPs denoting the ε4 alleles of *APOE*. The three isoforms of *APOE* are derived from combinations of the polymorphic SNPs rs429358 and rs7412^26^. The *APOE*-including GRS contains beta coefficients for the C alleles of both SNPs, defining ε4/ε4 genotype and *APOE4* isoform. Given the missingness of genotype data for some ALSPAC participants, GRSs were created for all individuals who had genotype data for at least one SNP for AD to preserve sample size and statistical power. The smallest number of SNPs for any included individual was 18 out of 25. These GRSs used the AD risk-increasing allele and log(OR) as external weights. Apart from *APOE* SNPs that were not carried forward to replication GWAS analyses, all effect sizes were obtained from the final stage of GWAS meta-analysis (*Supplementary Table 3*). The GRS reflects the average per-SNP effect on AD risk.

In UK Biobank, liability to AD was instrumented using the same SNPs used to create GRSs in ALSPAC. As such, the same data harmonisation process was used.

### Assessment of metabolites

In ALSPAC, blood samples were taken at clinics when offspring participants were approximately 8, 16, 18 and 25 years old. These samples were fasted except for those obtained at age 8 years. A total of 229 metabolites from a targeted metabolomics platform were measured via proton nuclear magnetic resonance (^1^H-NMR) spectroscopy using EDTA-plasma^27^. All metabolites were quantified at the first three time points; however, the following were not measured at 25 years: diacylglycerol, ratio of diacylglycerol to triglycerides, fatty acid chain length, degree of unsaturation, conjugated linoleic acid, and ratio of conjugated linoleic acid to total fatty acids.

Most metabolites relate to lipoproteins, categorised by density and size. Lipoprotein characteristics are recorded, including their triglyceride, phospholipid and cholesterol content. Various fatty acid, glycolysis-related, amino acid and inflammatory trait concentrations are also included.

In the UK Biobank, non-fasting EDTA plasma samples from a random subset of UK Biobank participants (n = 118 466) were analysed for levels of 249 metabolites, using the same platform as in ALSPAC, but with several additional ratios of lipid measures.

### Statistical approach

We adopted a “reverse MR”^28^ framework, such that genetic liability to AD is treated as the exposure and metabolites as the outcome, to ascertain the “metabolic features” of AD susceptibility in a preclinical population. *Figure 1* illustrates the potential mechanisms of association between AD SNPs and circulating metabolites. In ALSPAC, we conducted a GRS analysis which combines alleles into a score, whereas in the UK Biobank, we performed a formal MR analysis which uses SNPs as formal instrumental variables (IVs) for AD liability^29^. GRS analyses are typically better powered than MR analyses and hence were deemed more suitable for ALSPAC’s smaller sample size. GRS analyses do not, however, allow interrogation of potential bias due to horizontal pleiotropy. MR analyses are less well powered than GRS analyses, but there are several sensitivity analyses (including MR-Egger, weighted median and weighted mode^30^) that enable the assessment of horizontal pleiotropy.

**Figure 1.**
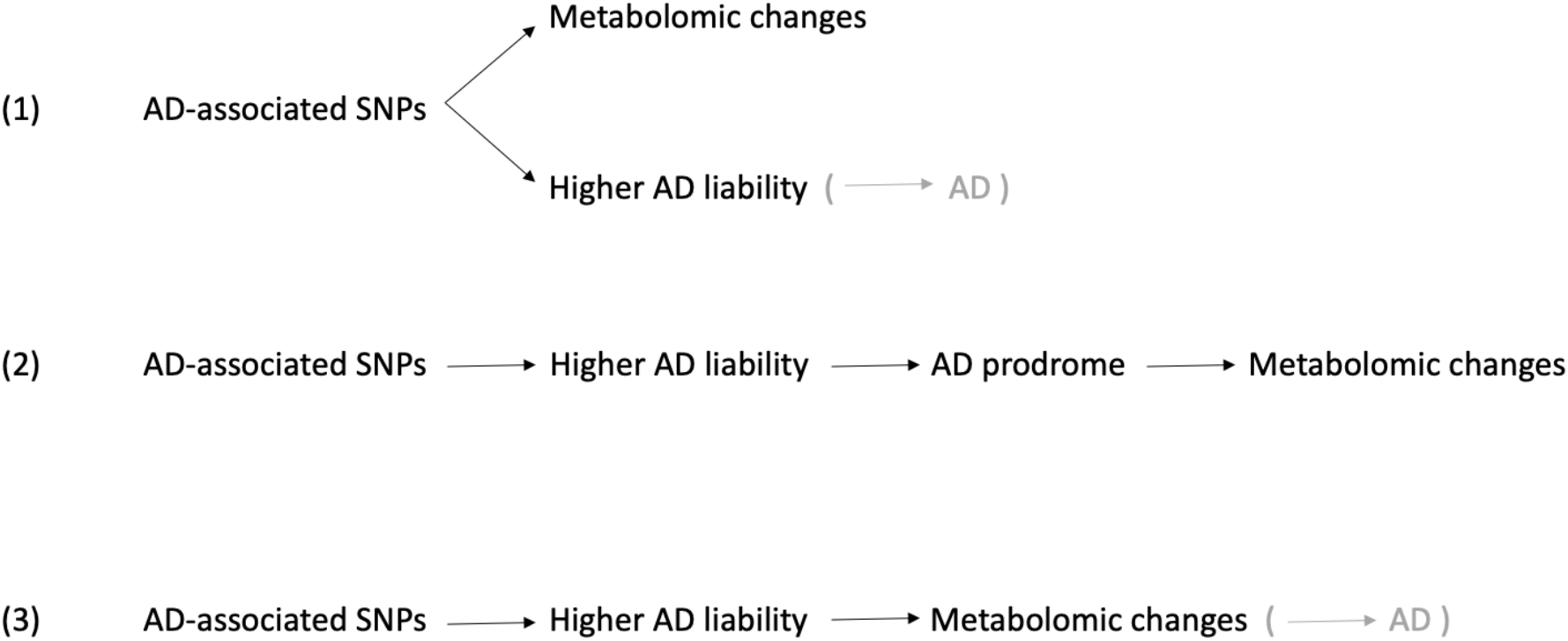
Schematic to demonstrate potential mechanisms of association between the AD GRS and metabolomic changes. Bracketed links denote hypothesised associations with AD itself, rather than our analysis (which does not directly measure AD but genetic liability to AD). (1) Horizontal pleiotropy. SNPs incurring higher AD liability exert their effects on AD via a different mechanism from their effects on metabolites. This is unlikely given the postulated role of metabolites in AD pathogenesis. However, APOE is known to be pleiotropic, performing location and isoform-specific effects. -= (2) Hallmarks of the AD prodrome causing metabolic derangements. This is unlikely as participants as young as those in ALSPAC would not be expected to be experiencing prodromal AD. (3) Here, metabolomic changes are a direct consequence of increased AD liability and precede the onset of clinical disease.

Results across cohorts are comparable despite the different analysis methods; all causal effect estimates were multiplied by 0.693 (loge2) as recommended and are interpreted as SD-unit differences in each metabolic trait per doubling of genetic liability to AD^31^.

In ALSPAC, the GRS and metabolite measurements were standardised by creating z-scores prior to analysis. Associations between the AD GRS and each metabolite at each time point were assessed using separate linear regression models, adjusting for age at time of metabolite assessment and sex.

In UK Biobank, 118,466 participants of European ancestry were stratified into tertiles of age, before a GWAS of metabolites was performed. Genetic association data for metabolites were generated using the MRC IEU UK Biobank GWAS pipeline^32^. All metabolites were standardized and normalized prior to analyses using rank-based inverse normal transformation. SNP-exposure associations based on the 25 SNPs for AD (the same SNPs used to create GRSs in ALSPAC) were integrated with the SNP-metabolite associations. Three statistical methods were used to generate MR estimates of effect using the TwoSampleMR package^33^: inverse variance weighted (IVW), MR Egger, weighted median, and weighted mode, each of which make differing assumptions about directional pleiotropy^34,35^. MR analyses were repeated with a set of 23 SNPs excluding the two *APOE4* SNPs. These analyses were performed in R version 4.0.2.

Summary-level GWAS results for metabolites can be accessed through the IEU-OpenGWAS platform^36^, accessible at https://gwas.mrcieu.ac.uk/datasets/?gwas_id__icontains=met-d.

## RESULTS

Full results for all associations between genetic liability to AD and metabolites, including and excluding *APOE* from analyses, can be found in Supplementary Tables 4, 5 and 6. Overall, when strong evidence was observed for causal effects of AD liability on metabolites (i.e., confidence intervals did not span the null), the direction and magnitude of association between genetic liability to AD (per SD higher GRS) and metabolite effect sizes remained consistent across the life course. Within ALSPAC, the extent of overlap of confidence intervals means it is often impossible to elucidate a trend in the associations across time points. The same is true for UK Biobank results, except for the main lipid metabolites, where there is a trend for attenuation of effect size towards the null as age increases in the IVW models. In general, UK Biobank estimates were largely consistent across MR sensitivity models, with wider confidence intervals for MR Egger estimates and smaller confidence intervals for weighted median and weighted mode estimates compared to IVW. For all metabolite subcategories, there is substantial attenuation of beta values towards the null, without substantial loss of statistical power when *APOE4* is excluded from the GRS (Supplementary Figures 2-4). This suggests that results are largely driven by the *APOE4* variant. Within ALSPAC, higher age was associated with wider confidence intervals, reflecting decreased sample size at each consecutive clinic visit.

### Genetic liability to AD and lipid traits

Of all metabolite subtypes, lipid traits demonstrated the strongest and most consistent associations with higher AD liability (including *APOE4*, Figure 2). The strongest positive associations were observed for the following lipid metabolites: serum total cholesterol, very-low density lipoprotein (VLDL) cholesterol, remnant cholesterol, low-density lipoprotein (LDL) cholesterol, esterified cholesterol, free cholesterol, apolipoprotein B and ratio of apolipoprotein B to apolipoprotein A1. For these metabolites, the magnitude and direction of association was similar across the life course. Within ALSPAC, the strongest positive associations tended to be observed at 18 years old (e.g., LDL cholesterol (0.11 SD; 95% CI 0.09, 0.14). For these same lipid metabolites, UK Biobank estimates from IVW models exhibited a decreased effect of AD liability with increased age tertile, although confidence intervals were overlapping (e.g., LDL cholesterol, youngest: 0.15 SD; 95% CI 0.07, 0.23, intermediate: 0.13 SD; 95% CI 0.07, 0.20, oldest: 0.10 SD; 95% CI 0.05, 0.16). Estimates from weighted mode and weighted median models showed similar differences between age tertiles for total, VLDL and LDL cholesterol and apolipoprotein B, but with non-overlapping confidence intervals between the intermediate and oldest tertiles. Across both cohorts, the effect of AD liability on high-density lipoprotein (HDL) cholesterol moved towards the null as age increased. There was no association with triglycerides in HDL at any ALSPAC time point, with confidence intervals consistently spanning the null. However, the effect estimates from IVW models for AD liability in UK Biobank were negative for triglycerides in HDL and decreased with age, which was consistent across sensitivity models. AD liability had no effect on apolipoprotein A1 but was consistently negative in UK Biobank. Effect estimates for the association between higher AD liability and sphingomyelins were consistently positive within ALSPAC (e.g., 25yrs: 0.07 SD; 95% CI 0.04, 0.09). This association persisted within each UK Biobank age tertile in IVW and sensitivity models. LDL triglycerides demonstrated increasing effects of AD liability with age in ALSPAC then decreasing effects with age in UK Biobank.

**Figure 2.**
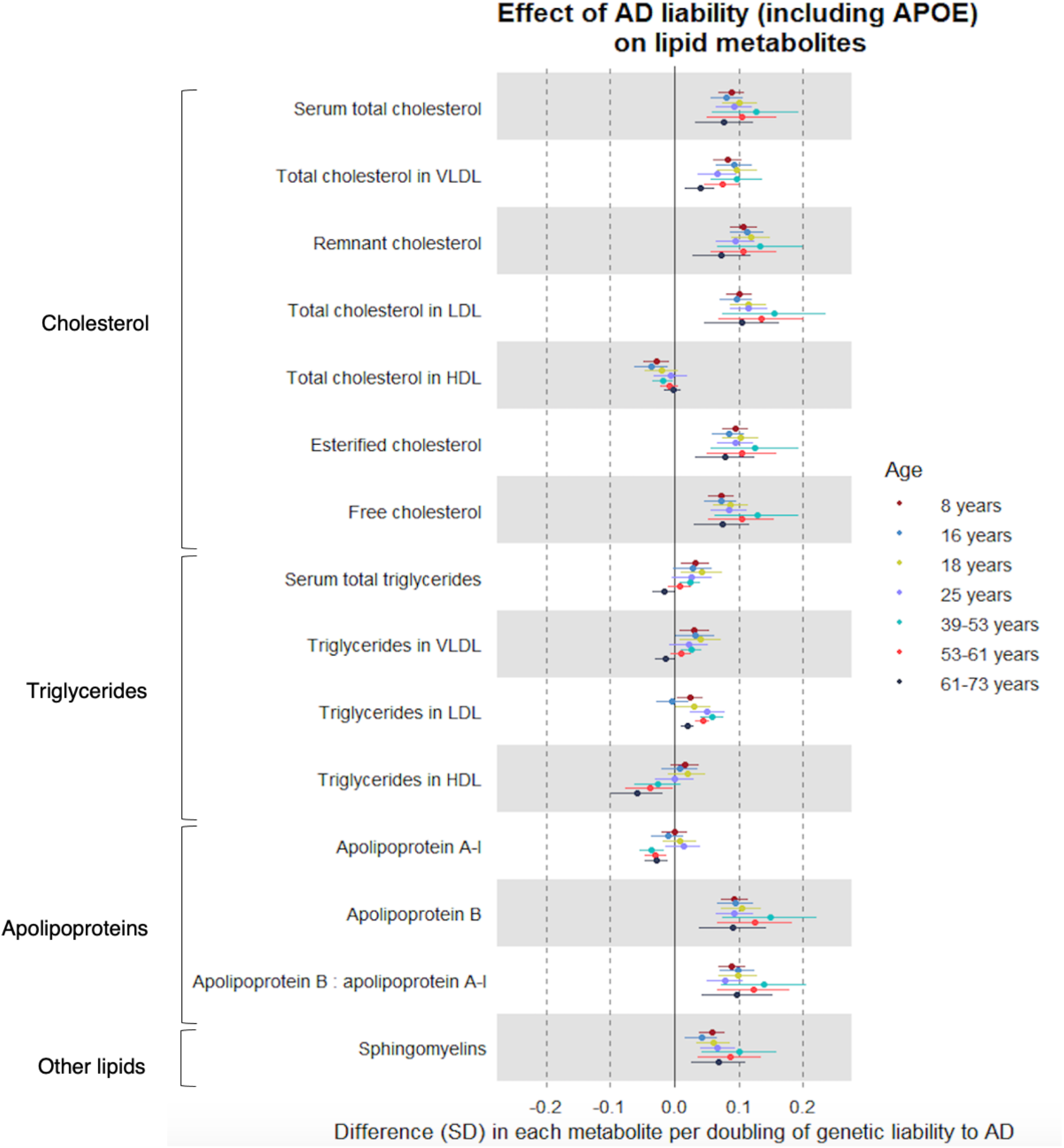
Forest plot showing the estimated effect of higher AD liability (including *APOE*) on main lipid metabolites. Effect size estimates for the first four time points are derived from ALSPAC (based on linear regression models). The latter three are from UK Biobank (based on IVW MR models), stratified by age (39-53 years, 53-61 years, 61-73 years).

### Genetic liability to AD and fatty acids

When including *APOE* in the GRS, higher AD liability had a strong positive association with many fatty acid (FA) metabolites and an attenuated association with corresponding fatty acid ratios. The FA metabolites most strongly associated with higher AD liability were total FA, linoleic acid, omega-3 FA, omega-6 FA, polyunsaturated FA, monounsaturated FA, and saturated FA (Figure 3). Higher AD liability was associated with an increase in each of these FA metabolites at all seven time points, suggesting that these associations persist across the life course. Within ALSPAC, for these FA metabolites, as was observed with lipids, the strongest positive association was at 18 years old (e.g., total FA at 18yrs, 0.07 SD; 95% CI 0.04, 0.10), except omega-3 FA (strongest ALSPAC association at 25yrs (0.06 SD; 95% CI 0.03, 0.08)). In the oldest UK Biobank tertile, the effect of AD liability on these main FA estimates was decreased compared with the youngest tertile, consistently across models (IVW: e.g., total FAs, oldest: 0.02 SD, 95% CI 0.01, 0.03, youngest: 0.06 SD, 95% CI 0.04, 0.08).

**Figure 3.**
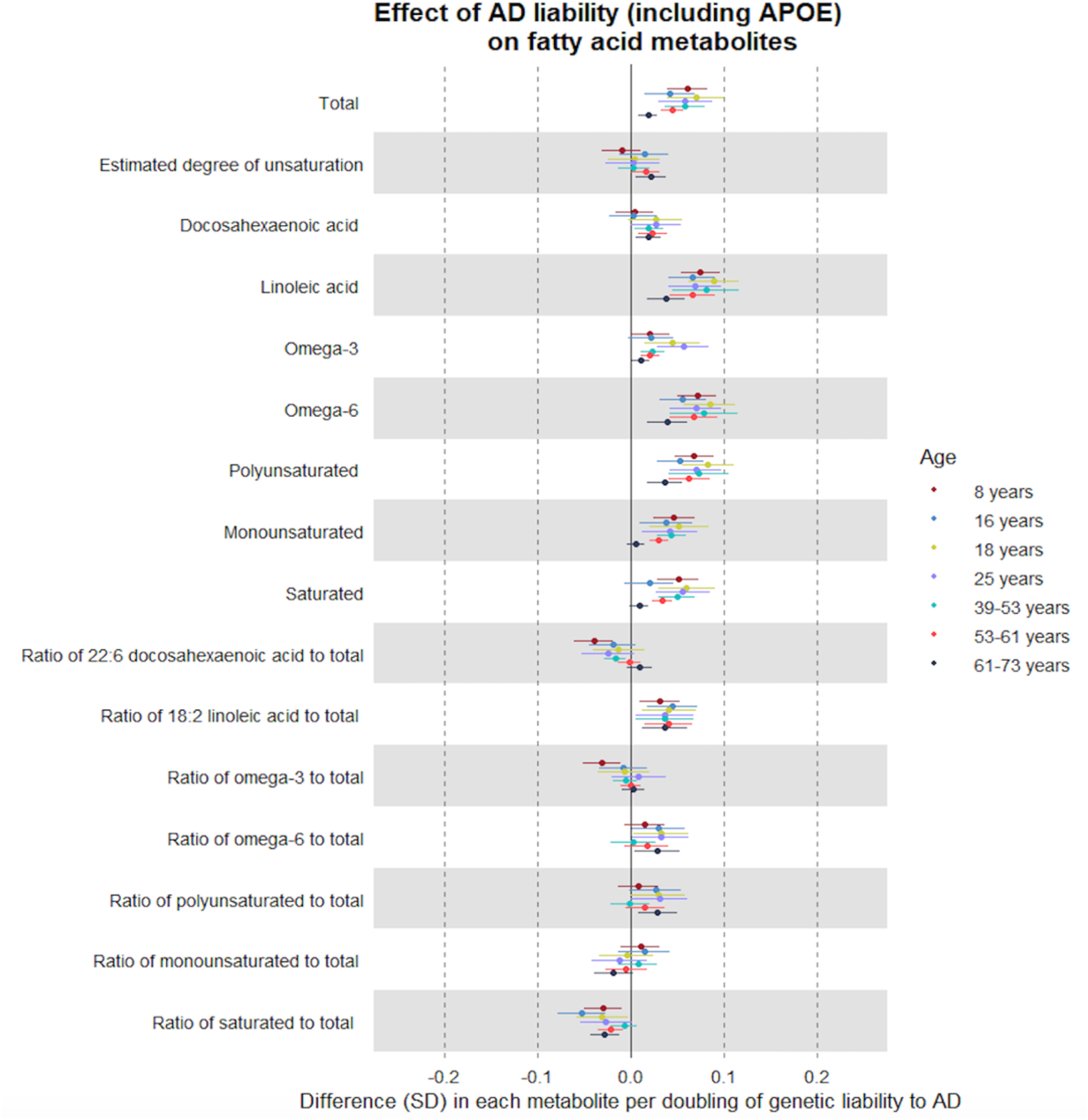
Forest plot showing the estimated effect of higher AD liability (including *APOE*) on fatty acid metabolites. Effect size estimates for the first four time points are derived from ALSPAC (based on linear regression models). The latter three are from UK Biobank (based on IVW MR models), stratified by age (39-53 years, 53-61 years, 61-73 years).

The effect of liability to AD on the ratio of docosahexaenoic acid (DHA) to total FAs turned from negative to null with increasing age group across both cohorts, whilst the effect on the ratio of linoleic acid to total FAs remained consistent across all age groups.

### Genetic liability to AD and non-lipid traits

#### Glycolysis-related traits

The three glycolysis-related traits included in this platform are glucose, citrate, and lactate. In ALSPAC, effect sizes for these metabolites centre around zero, though the imprecision of confidence intervals means that the magnitude and direction of potential effects is unclear. In UK Biobank, IVW effect estimates tended to be more precise than ALSPAC for these traits, though they still largely crossed the null. Within the oldest UK Biobank tertile, all models showed an inverse effect of AD liability on citrate. In ALSPAC, associations of AD liability with lactate were more positive at older ages (25yrs: 0.04 SD; 95% CI 0.01, 0.06), but the effect estimates for lactate in all UK Biobank age tertiles were negative (e.g., IVW, oldest: -0.01 SD; 95% CI -0.02, 0.00).

#### Amino acids and inflammation

Of all metabolite subcategories, amino acids (including the branched chain amino acids (BCAAs) isoleucine, leucine, and valine) demonstrated the weakest association with higher AD liability including *APOE* (Figure 4). There were no consistent positive associations with any amino acids at any time point in ALSPAC. At 16yrs, increased AD susceptibility was associated with slightly decreased levels of leucine (−0.03 SD; 95% CI -0.05, -0.005), valine (−0.03 SD; 95% CI -0.05, -0.01) and tyrosine (−0.03 SD; 95% CI -0.06, -0.004). In UK Biobank, liability to AD had an inverse association with several amino acids (e.g., tyrosine, youngest tertile: -0.02 SD; 95% CI -0.03, 0.00). There was no association of higher AD liability with glycoprotein acetyls, a marker of inflammation, at any time point in ALSPAC or UK Biobank IVW models. However, estimates from weighted median and weighted mode models demonstrated weak positive associations of AD liability and glycoprotein acetyls in the youngest and intermediate UK Biobank tertiles (Supplementary Tables 8 and 9).

**Figure 4.**
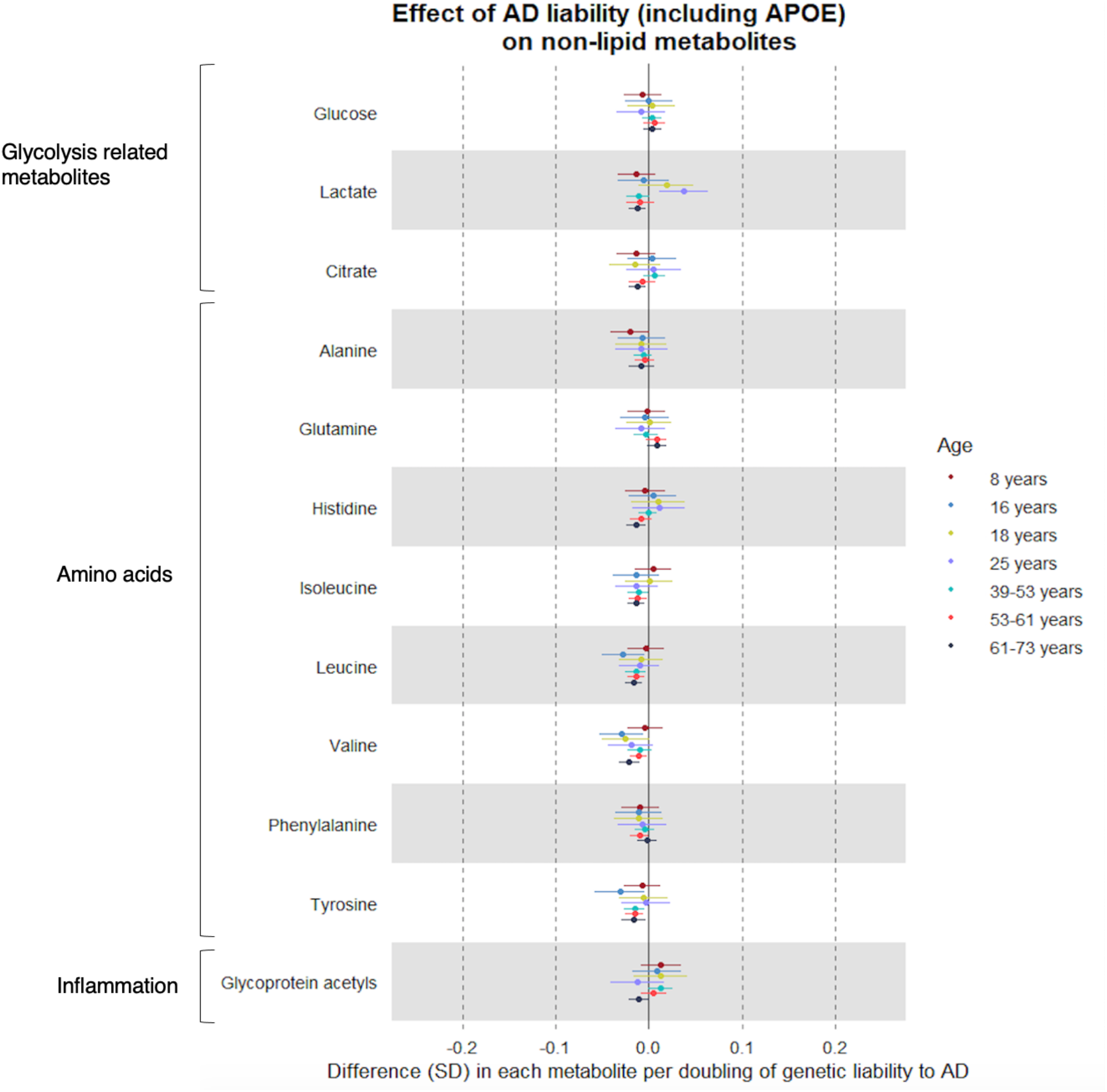
Forest plot showing the estimated effect of higher AD liability (including *APOE*) on glycolysis-related, amino acid and inflammatory traits. Effect size estimates for the first four time points are derived from ALSPAC (based on linear regression models). The latter three are from UK Biobank (based on IVW MR models), stratified by age (39-53 years, 53-61 years, 61-73 years).

## DISCUSSION

This study aimed to estimate the effects of genetic liability to AD on the circulating metabolome measured across early life and into adulthood, revealing potential early stages of AD pathophysiology. Prior studies were limited in their ability to determine whether metabolic perturbations were a cause or consequence of disease activity. However, given the young age of ALSPAC participants, we are confident that observed effects in this cohort precede clinical AD and are therefore not consequences of AD pathophysiology. Our most striking finding is the profound and enduring influence of the *APOE4* isoform on lipid and fatty acid traits, which was evident already in childhood and persistent into later adulthood. There were notably null associations of AD liability with glycolysis- and inflammatory-related traits, suggesting that AD liability is more specifically reflected in lipid metabolism, driven by *APOE*.

It has been hypothesised that the association of higher AD liability (including *APOE4*) with lipid traits is via its effect on atherosclerosis. This is supported by both comparable enrichment of plasma lipid subtypes for AD and cardiovascular disease^37^ and demonstration here of the strongest positive associations being for the inherently proatherogenic traits LDL cholesterol, apolipoprotein B and ratio of apolipoprotein B to apolipoprotein A-1^38^. It has also been shown that elevated LDL cholesterol is associated with increased cerebral amyloid deposition^39^. This may constitute an additional pathophysiological mechanism of lipids in AD, particularly in *APOE4* carriers.

We found evidence that associations of HDL-C and its major constituent apolipoprotein A-1 with higher AD liability were weakly negative. Observationally, and in MR studies, higher HDL-C is associated with lower AD risk^40,41^. Results from a previous prospective cohort study implicated lower plasma apolipoprotein A-1 with elevated risk of AD progression in ε4 carriers^42^. That study was limited by modest sample sizes and relatively short follow-up (mean 2.5±1.6 years). This study compliments their findings as the strength of association of apolipoprotein A-1 with AD liability increases across the life course.

Our study found a weak positive association between higher AD liability and sphingomyelins levels that was consistent across age groups. Sphingolipids are a class of lipids, of which sphingomyelins are members^43^. Study of post-mortem brains, CSF and plasma have implicated sphingomyelin perturbations in AD pathophysiology^44^, though prospective results are inconsistent. A recent targeted metabolomics study of blood and brain found, however, that increased SM levels correlated with AD severity, tracking disease progression from prodromal to preclinical stages^45^.

We show that the effect of higher AD liability (including *APOE4*) on triglyceride levels in VLDL, HDL and total triglycerides weakens with age, which could reflect increased medication use with age (e.g., statins) or that these are metabolic signatures of prodromal AD but not the disease itself. The magnitude of causal associations are considerably less than what was observed for a recent untargeted lipid profiling study by Bernath et al. concluded that AD-mediated effects on triglycerides were specific to carriers of the ε4 allele^46^, supporting the results of this study. Elevated triglycerides in VLDL precede amyloid deposition in mouse models^47^, such amyloidosis being a hallmark of AD neuropathology. These results therefore exemplify the substantial effect of the *APOE4* isoform on lipid metabolites.

Except for several fatty acid traits, the associations between higher AD liability and other non-lipid metabolites were broadly null in our study. When including *APOE4* in the GRS, strong positive associations were observed with total FAs, linoleic acid (LA), omega-6 FAs and polyunsaturated FAs. Corresponding FA ratios, which may better reflect FA biology^27^, demonstrated an attenuated, yet still positive association with higher AD liability. Aside from functioning as membrane constituents and energy sources^48^, FAs mediate inflammation^49^, a process central to the pathogenesis of both CVD and AD^50,51^. LA has previously been associated with the extent of AD neuropathology in a nontargeted metabolomics study^52^, though small sample size and confounding limit causal inference.

As with lipid metabolites, the association between fatty acid concentrations and higher AD liability appears to be mediated by *APOE* – without this locus, all associations attenuated towards the null. Regarding clinical implications, we must be mindful that given the central-peripheral flux of FAs^45^, it is difficult to elucidate whether these peripheral/blood markers are representative of central/brain pathology. As such, future work should compare plasma metabolites with those in CSF and brain tissue.

Associations between AD liability and glycolysis-related traits were generally null. Type 2 diabetes, defined as elevated plasma glucose, is hypothesised to be a risk factor for AD^53^, although MR studies to date have not supported any causal association^54–56^. Diabetes mechanisms mediate the pathological effects of the ε4 genotype^57^ and influence cerebral glucose metabolism^58^. Results from a prospective cohort study with several decades of follow-up additionally demonstrate that plasma glucose derangements are only evident in ε4 carriers from midlife onwards^59^. However, even in the oldest UK Biobank tertile, we observed little evidence of effect of AD liability on glucose.

The association of AD liability with lactate was positive at older ages in ALSPAC, but inverse or null in UK Biobank age tertiles. Increased lactate in the CSF and brain has been associated with higher AD risk, the degree of perturbation correlating with extent of neurodegeneration^60^. *In vitro* evidence suggests that this trend may be ε4-mediated^61^, resulting from a ‘Warburg like’ endophenotype that is present in humans many decades before onset of AD^62^. The lack of consistency of effect of AD liability on lactate levels across different life-stages limits its clinical utility as a biomarker of early disease.

Evidence for the role of BCAAs in AD is inconclusive. Observationally, increased BCAA levels appear to protect against AD^63^, which is supported by our inverse effect estimates for AD liability on BCAAs in UK Biobank. Increased BCAAs have, however, been robustly associated with both increased insulin resistance and diabetes risk in MR analysis^64,65^. The absence of association between increased BCAAs and higher AD liability in the present study perhaps suggests that the link between BCAAs and AD is mechanistically distinct from pathways of glucose and insulin metabolism. This, however, contradicts results from a recent MR analysis that concluded those predisposed to raised plasma isoleucine levels are at an increased rather than decreased risk of AD^66^.

Our analyses are underpinned by three core IV assumptions that must be satisfied for results to be valid. The first assumption of robust association between the IV and trait of interest was fulfilled given the large GWAS sample size and inclusion of SNPs relating to genes with known *a priori* biological function in relation to AD (*APOE*). The second assumption is that of no confounders of the IV and the outcome. This was addressed to the extent possible here by using a largely ancestrally homogenous population (>96% white ethnicity). The final assumption is that there is no association of genetic instruments with the outcome, except via the exposure of interest. Our UK Biobank results demonstrated consistency across pleiotropy-robust models, indicating that horizontal pleiotropy is unlikely to be causing the associations we see.

## STUDY LIMITATIONS

A key limitation of this study is the modest analysis sample size for ALSPAC analyses, particularly at older ages, however our use of an allele score method did appear to enable relatively high statistical power and precision of exposure-outcome estimates. The lack of ancestral diversity in ALSPAC (96% white) and UK Biobank (only Europeans analysed) limits the generalizability of results to diverse populations, but does, however, limit the potential for confounding by population stratification. Given that the number of ε4 alleles is a stronger predictor of AD for those of European ancestry than those of African American or Hispanic ancestry^67^, future studies must investigate the extent to which *APOE4* carrier status influences the metabolome for other populations. Despite the central-peripheral flux of metabolites via the blood-brain barrier, previous studies have noted that the AD molecular profiles of plasma and CSF are divergent^68^. Therefore, the extent to which inferences regarding central AD pathophysiology can be made from this study are potentially limited. Future work should therefore compare the effect of higher AD liability on plasma and CSF metabolites, although such data do not yet exist at scale. UK Biobank and the ALSPAC 8-year metabolite measurements were taken from nonfasted blood samples, whilst samples from all other timepoints were fasted, which potentially limits the comparability of UK Biobank and age 8 with the other ages. Another limitation is the targeted nature of the Nightingale metabolomics platform, which focuses on metabolites previously identified to be of clinical interest, most of which are lipids. An untargeted approach would allow discovery of unknown biomarkers, including those beyond the lipid classes, of AD liability.

## CONCLUSIONS

The results of this study support a profound effect of *APOE4* in producing an early metabolic signature of higher AD liability that persists throughout the life course and demonstrates that other AD risk variants have minimal effects of circulating metabolites. This signature constitutes elevated proatherogenic lipid traits, namely LDL cholesterol and its major protein, apolipoprotein B, in addition to sphingomyelins. *APOE4*-mediated effects are also observed for fatty acid traits, in particular omega-6 FAs and linoleic acid. These AD-associated metabolic derangements begin in childhood, many decades before the emergence of disease, and persist into later adulthood when AD is more commonly diagnosed.

## Data Availability

Individual-level ALSPAC data are available following application. This process of managed access is detailed at www.bristol.ac.uk/alspac/researchers/access. Summary-level GWAS results can be accessed through the IEU-OpenGWAS platform, accessible at https://gwas.mrcieu.ac.uk/datasets/?gwas_id__icontains=met-d.

## Acknowledgements

We are extremely grateful to all the families who took part in this study, the midwives for their help in recruiting them, and the whole ALSPAC team, which includes interviewers, computer and laboratory technicians, clerical workers, research scientists, volunteers, managers, receptionists and nurses. This work has arisen from a research project undertaken by Hannah Compton as part of their iBSc in Genomic Medicine at Bristol Medical School.

## SUPPLEMENTARY MATERIAL

Supplementary Tables 1-9 are in an excel file.

**Supplementary Figure 1.**
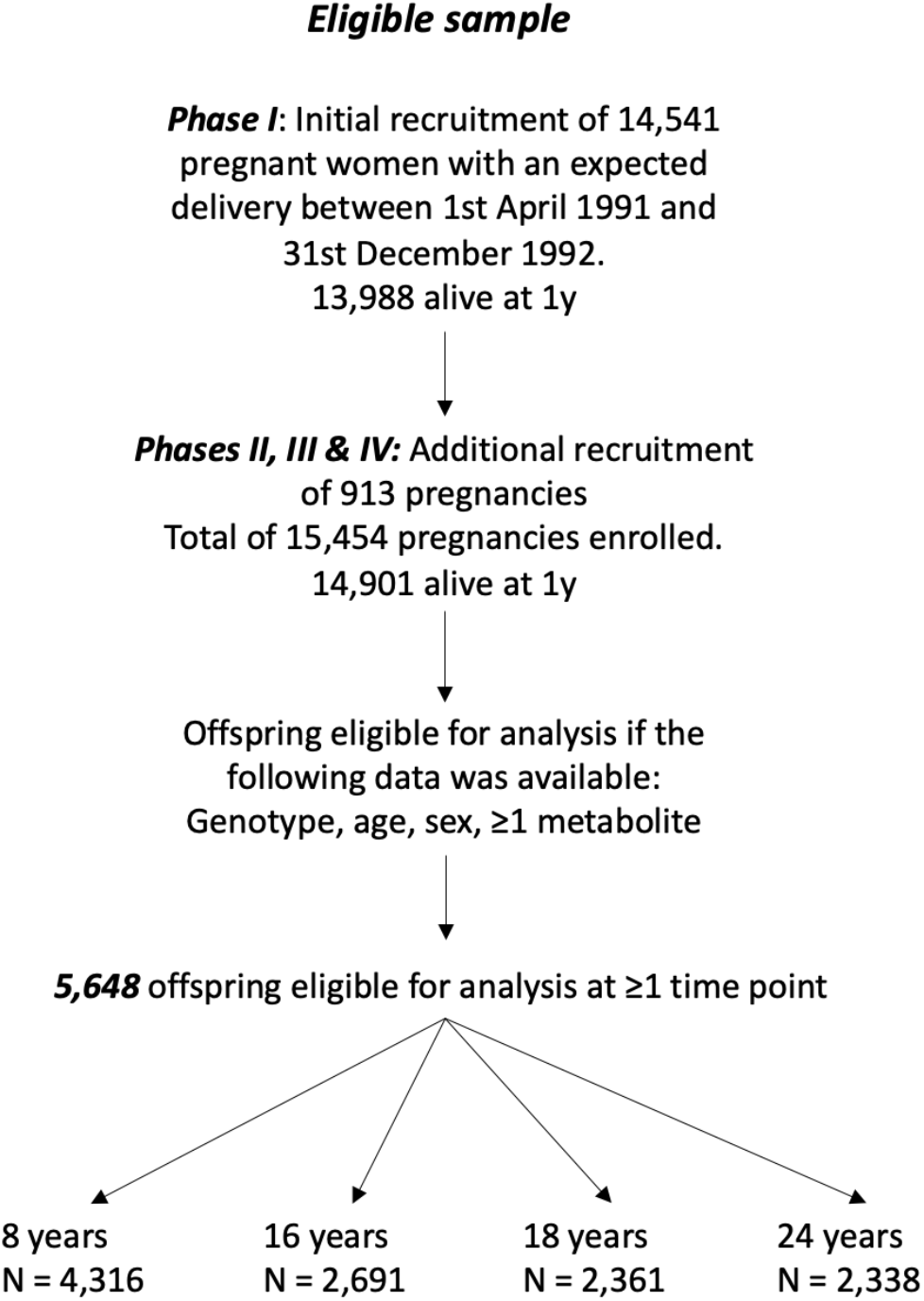
Flow chart to demonstrate eligibility criteria for inclusion to ALSPAC analyses

**Supplementary Figure 2.**
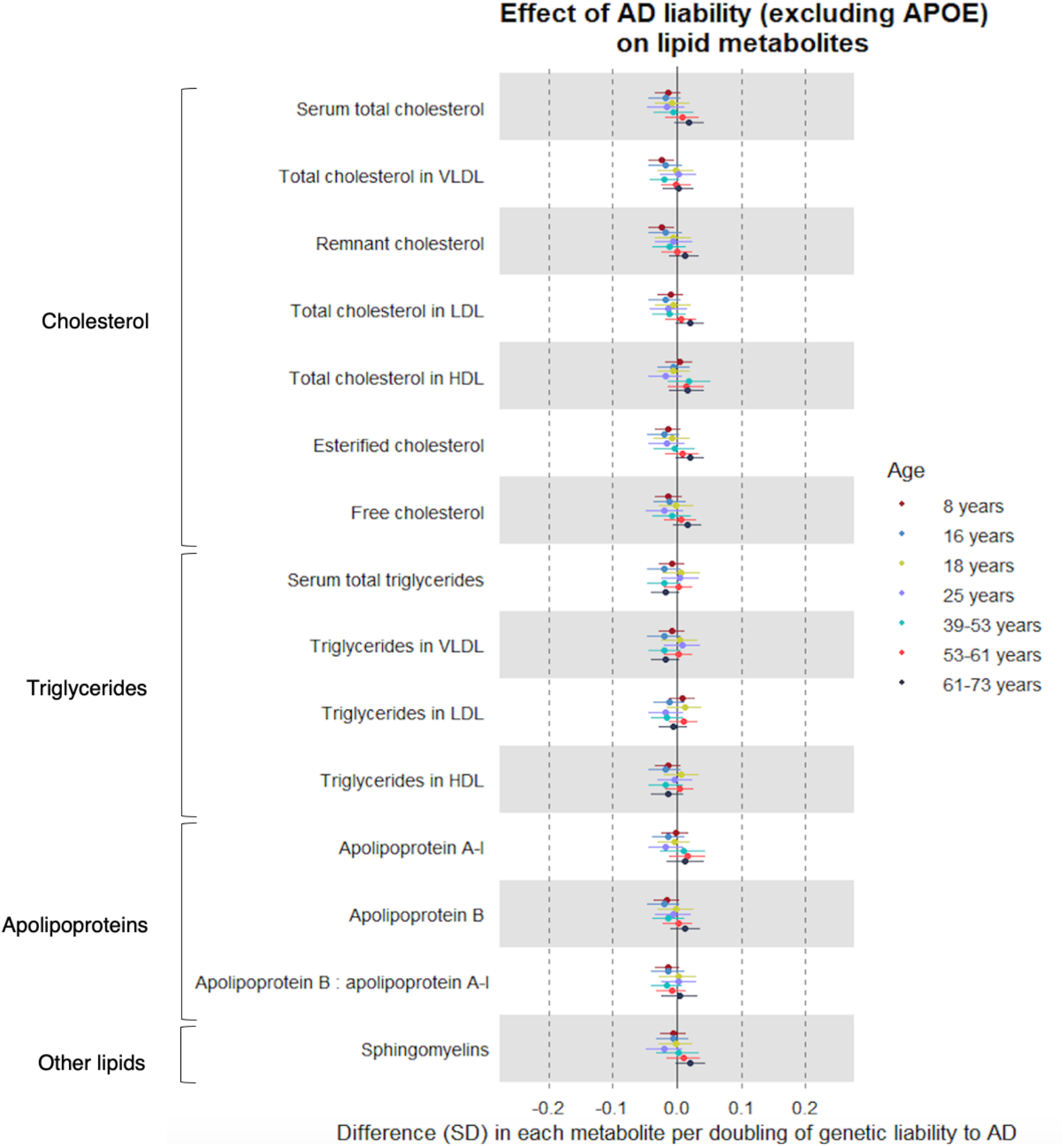
Forest plot showing the estimated effect of higher AD liability (excluding *APOE*) on lipid metabolites. Effect size estimates for the first four time points are derived from ALSPAC (based on linear regression models). The latter three are from UK Biobank (based on IVW MR models), stratified by age (39-53 years, 53-61 years, 61-73 years).

**Supplementary Figure 3.**
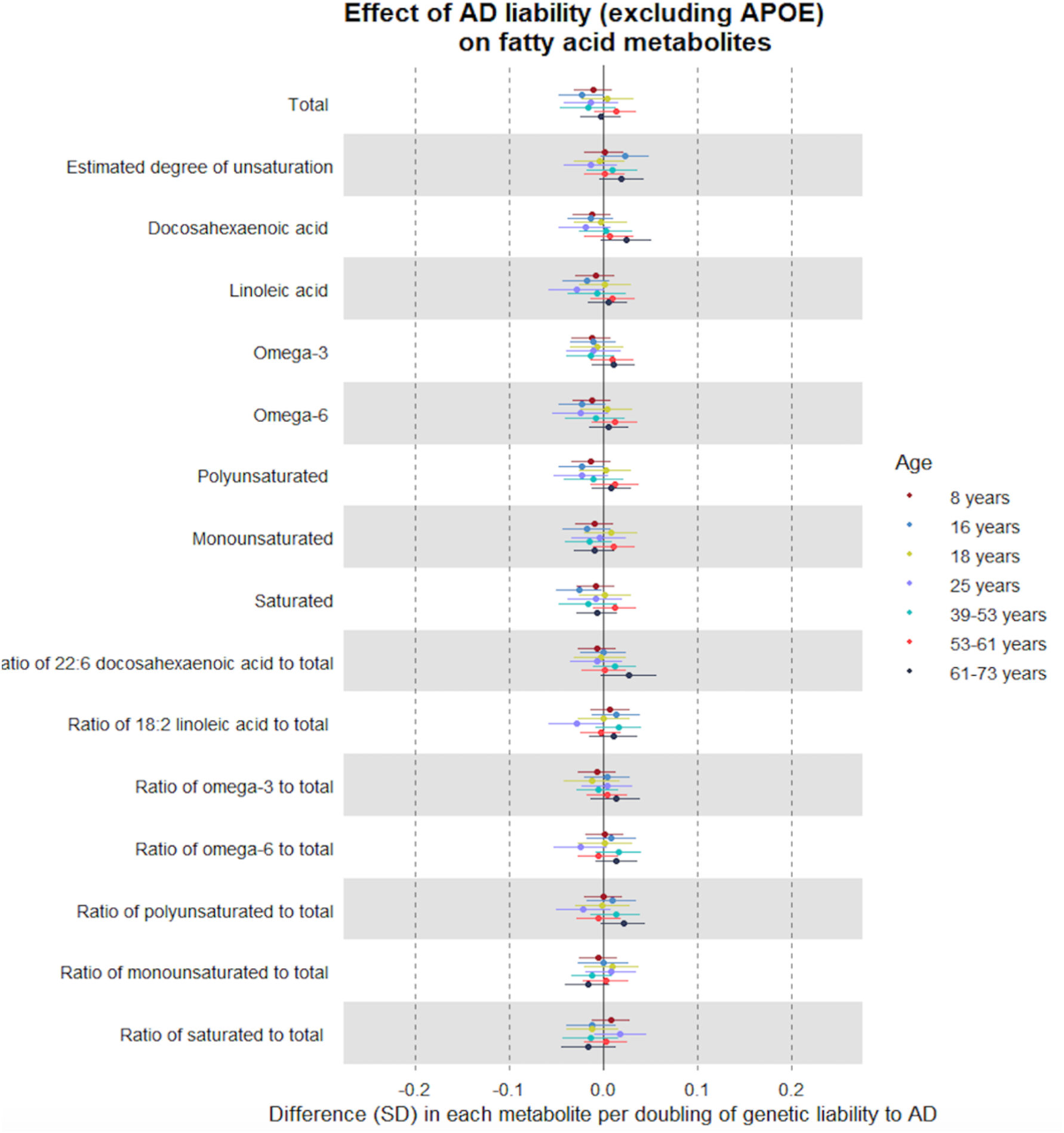
Forest plot showing the estimated effect of higher AD liability (excluding *APOE*) on fatty acid metabolites. Effect size estimates for the first four time points are derived from ALSPAC (based on linear regression models). The latter three are from UK Biobank (based on IVW MR models), stratified by age (39-53 years, 53-61 years, 61-73 years).

**Supplementary Figure 4.**
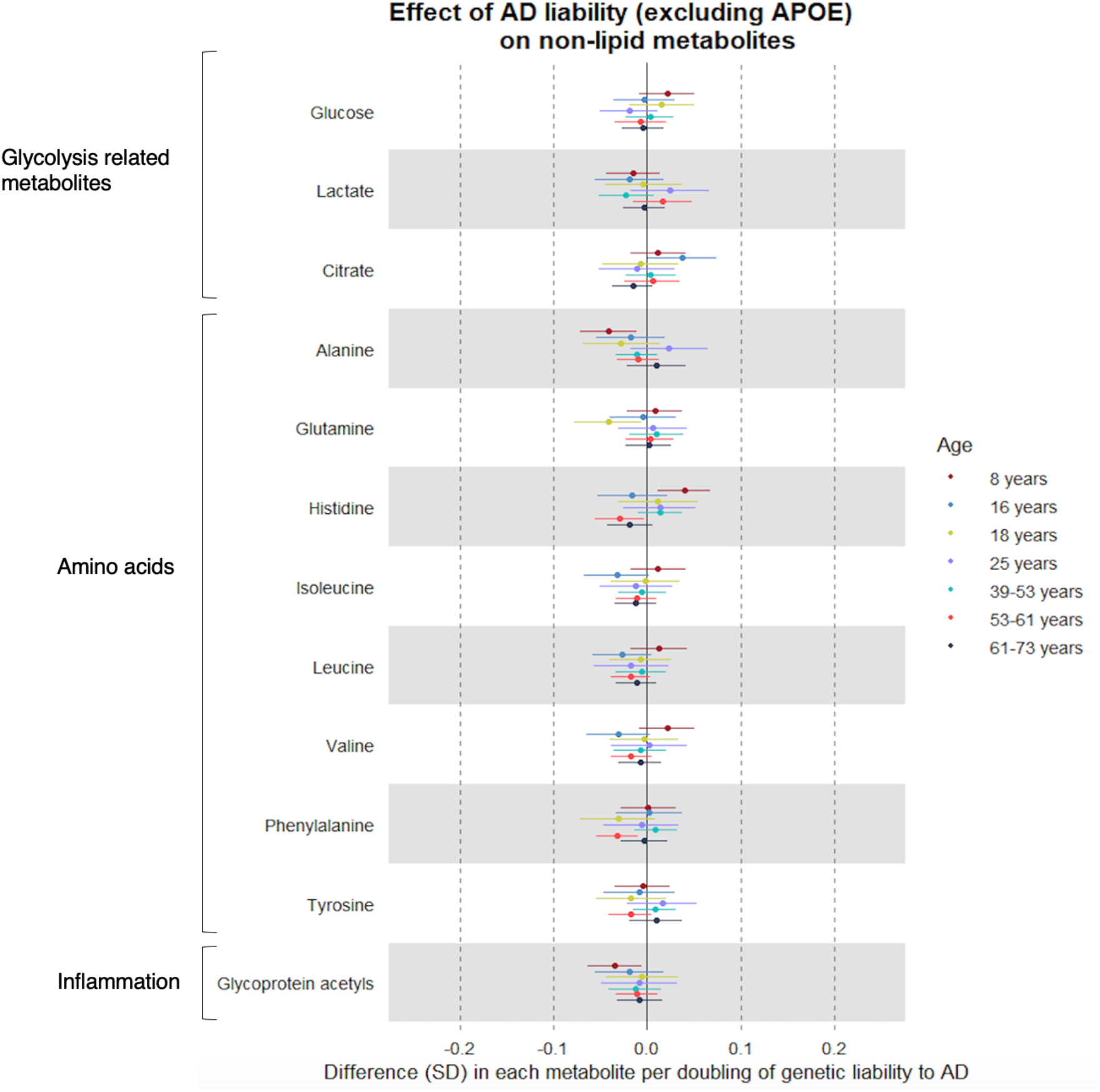
Forest plot showing the estimated effect of higher AD liability (excluding *APOE*) on glycolysis-related, amino acid and inflammatory traits. Effect size estimates for the first four time points are derived from ALSPAC (based on linear regression models). The latter three are from UK Biobank (based on IVW MR models), stratified by age (39-53 years, 53-61 years, 61-73 years).

